# *TG-Interacting Factor 1* expression quantitatively impacts survival in acute myeloid leukemia

**DOI:** 10.1101/2020.02.04.20020537

**Authors:** Ling Yan, Julie A. Means-Powell, Danko Martincic, Vladimir D. Kravtsov, Yu Shyr, John P. Greer, Utpal P. Davé, Mark J. Koury, David Wotton, Rizwan Hamid, Stephen J. Brandt

## Abstract

Applying transcriptional profiling analysis to myeloblasts from 59 adult patients with acute myeloid leukemia (AML) treated at our institution, we found that expression of the three-amino acid loop extension (TALE) homeobox gene *TG-Interacting Factor 1* (*TGIF1*) correlated with overall and relapse-free survival, which was then confirmed in two other cohorts of patients.

Moreover, *TGIF1* expression correlated with survival for all cytogenetic risk groups and was an independent prognostic factor in multivariate analysis. To elucidate the mechanism, we used *Tgif1* knockout mice in which acute or chronic myeloid leukemia was induced through retroviral transfer of the *MLL-AF9* or *BCR-ABL* fusion genes into bone marrow cells. Loss of *Tgif1* accelerated disease progression, shortened survival, attenuated the response to chemotherapy, and doubled the frequency of leukemia-initiating cells. RNA-based sequencing analysis showed that genes associated with transforming growth factor-β (TGF-β) and retinoic acid signaling pathways were differentially affected in *Tgif1*^-/-^ compared to *Tgif1*^*+/+*^ leukemia cells.

## Introduction

Acute myeloid leukemia (AML) is an aggressive and heterogeneous disease that most commonly occurs in the elderly. Although a majority of patients achieve a complete remission (CR) with induction and consolidation chemotherapy, most will eventually relapse, with overall survival (OS) ranging from 10-35% *^1, 2^*. As a result, clinical indicators have been sought that could inform post-remission therapy and improve long-term survival *^3, 4^*. Established predictors of outcome include patient age, white blood cell (WBC) count, antecedent myelodysplastic syndrome, and, of greatest significance, the presence of specific chromosomal rearrangements in leukemia cells *^5^*. Patients with t(15;17), t(8;21), or inv(16) enjoy the longest overall survival (OS), while t(6;9), 11q23, monosomy of chromosomes 5 or 7, and complex karyotypes confer the worst prognoses, with 80% of these individuals ultimately relapsing *^6^*. The largest group of patients, however, has either a normal karyotype or specific cytogenetic abnormalities that put them at some intermediate risk of relapse and death *^7, 8^*.

Patients in the favorable risk group can be successfully treated with intensive chemotherapy only *^8^*, while those in the high-risk group require an allogeneic hematopoietic stem cell transplant (SCT) in remission for long term survival *^9^*. In contrast, the optimum therapy for intermediate-risk patients is unclear. Given the substantial number of individuals in whom a cytogenetic marker is not present or prognostic, additional, especially molecular, predictors of survival have been sought. One of the most extensively characterized molecular abnormalities is an internal tandem duplication (ITD) of the gene for Fms-Related Tyrosine Kinase 3 (FLT3). While found to impact survival in some analyses *^10, 11^*, no difference in that of *FLT3* ITD-positive and ITD-negative patients was detected in other studies *^12, 13^* as assessed independently of other mutations, in particular those in the *nucleophosmin* (*NPM1*) gene. More than one ITD *^10^*, a high ratio of mutant to wild-type DNA *^12, 13^*, and loss of the wild-type allele have been more clearly linked to a poor prognosis.

Rearrangement and/or altered expression of other genes altered or not have been found to have prognostic value. Increased expression of the oncogene *Ecotopic Viral Integration Site 1* (*EVI1*), for example, was associated with inferior survival in adults with *de novo* AML *^14^*, while reduced expression of the *Deleted in Colorectal Carcinoma* (*DCC*) gene was an independent prognostic factor for CR and OS *^15^*. Further, overexpression of the *Brain And Acute Leukemia, Cytoplasmic* (*BAALC*) gene had an adverse impact on overall survival (OS) and disease free survival (DFS) in a study of younger adults with cytogenetically normal *de novo* AML *^16^*, and mutation of the gene encoding the myeloid transcription factor C/EBP*α* (*CEBPA*) was associated with a better outcome in patients in the intermediate-risk group *^17, 18^*. Increasingly, expression profiling and deep sequencing approaches have been used to distinguish groups of patients differing in biology as well as survival.

Analyzing global gene expression profiles from a cohort of patients treated at our institution, we discovered that the abundance of mRNA for the three-amino acid loop extension (TALE) superclass homeobox gene *TG-Interacting Factor1* (*TGIF1*) *^19-22^* correlated very significantly with measures of survival. Analysis of its actions in *Tgif1* knockout mice in which acute or chronic myeloid leukemia was induced revealed that *Tgif1* abundance affected the number and/or function of leukemia-initiating cells (LICs), also referred to as leukemia stem cells (LSCs). This observation has important prognostic and therapeutic implications.

## Results

### TGIF1 expression is associated with clinical outcomes in AML

To identify new predictors of outcome, we carried out microarray analysis of leukemia cellular RNA from 59 of the 60 adult patients with AML treated at our institution (Supplemental Table S1). Modified Cox regression analysis was then used to identify genes whose expression correlated with OS. A group of 38 genes were predictive with a P < 0.001 and of 5 genes with a P < 0.0001, while the expression of a single gene, *TGIF1*, correlated with OS with the greatest significance (P < 0.00001).

Kaplan-Meier analysis of patient survival in this cohort showed that only 8% (3/39) of individuals whose leukemia cells contained lower levels of *TGIF1* mRNA than the reference remained alive at last follow-up (median survival = 181 days, confidence interval (CI) = 112-249 days), while 60% (12/20) of patients who expressed *TGIF1* at higher levels survived. Since more than half of patients with higher *TGIF1* expression were living at last analysis, median survival could not be determined. Log-rank analysis revealed that individuals with lower *TGIF1* expression had significantly shorter OS (Figure 1A) and relapse-free survival (RFS) (Figure 1B) than the group with higher expression. Although a commercial RNA preparation was used to stratify patients into lower and higher expressers, the best cut-point in *TGIF1* expression for discriminating survival determined by receiver operating characteristic analysis was close to that in the commercial reference (data not shown). When patients were segregated by *TGIF1* expression in quartiles, a concentration-dependent relationship between *TGIF1* expression and outcome was suggested (Figure 2A). Finally, *TGIF1* expression correlated with OS with even greater statistical significance when analyzed as a continuous variable (P < 0.0001).

**Figure 1.**
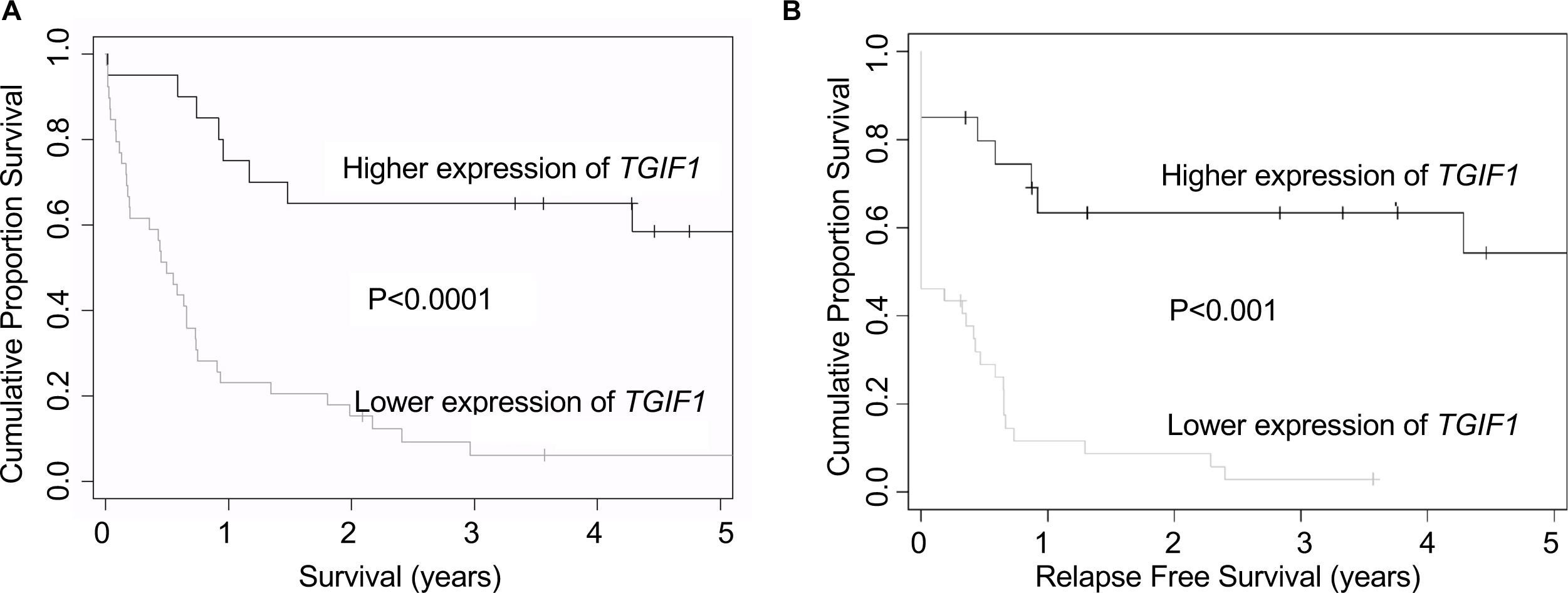
Lower *TGIF1* expression was associated with adverse and higher *TGIF1* expression associated with favorable clinical outcome in AML. **A**, OS in patients (n = 20) with higher expression relative to reference is shown in top curve. Median survival in this group was not reached at last follow-up, with 70% of this group still alive. The survival of patients (n = 39) with lower expression is shown in bottom curve. Median survival in this group was 181 days (CI = 112-249 days), with 7.6% still alive. **B**, RFS in patients (n = 20) with *TGIF1* expression greater than reference is shown in top curve. The survival of patients (n = 39) with lower *TGIF1* expression is shown in bottom curve.

**Figure 2.**
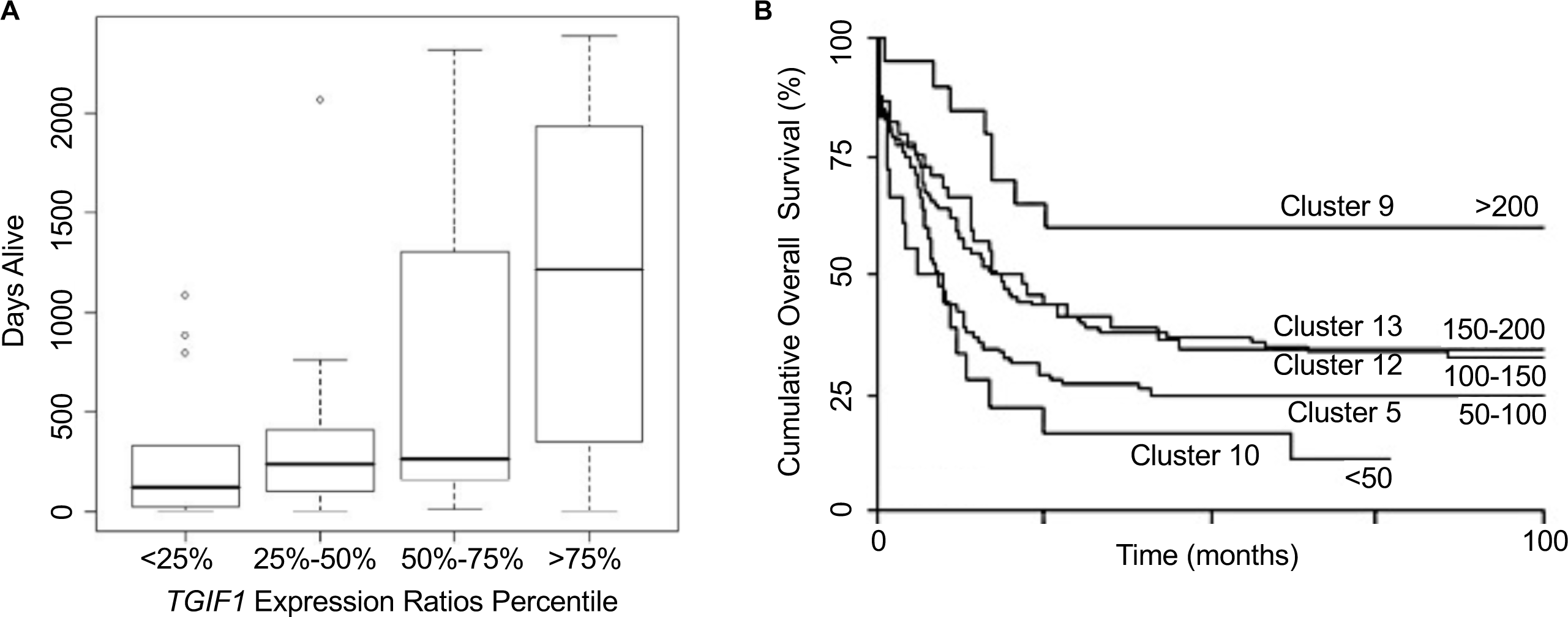
Analysis of OS in patients as a function of *TGIF1* mRNA expression. **A**, Box-and-whiskers plot of OS expressed as days alive vs. ratio of *TGIF1* expression in leukemia cells to commercial reference. Cases were subdivided into 4 groups according to *TGIF1* expression ratios as shown. **B**, *TGIF1* expression determined in 285 AML patients with Affymetrix U133A GeneChips (probe set 203313_s_at) and cases subdivided according to *TGIF1* expression into 5 groups: < 50 (n = 20), 51-100 (n = 45), 101-150 (n = 107), 151-200 (n = 95), and > 200 (n = 18). Log-rank analysis was carried out using Microsoft Excel software.

### TGIF1 expression correlated with survival for risk groups defined by cytogenetics analysis

Seventeen percent (1/6) and 86% (18/21) of patients with good- and poor-risk cytogenetic findings, respectively, had lower *TGIF1* expression. In contrast, *TGIF1* expression was more evenly distributed in patients with a normal karyotype. Specifically, 41% (11/27) had higher and 59% (16/27) lower *TGIF1* mRNA than reference, and individuals in the intermediate-risk group with lower *TGIF1* survived for significantly shorter times (median survival = 201 days, CI = 138-263 days) than those with higher expression (median survival = 1566 days). *TGIF1* expression also correlated with shorter OS for patients in poor-risk patients (data not shown) and even for the good-risk group, although the number of patients was small. Thus, *TGIF1* levels correlated with clinical outcome in adult AML patients from all three cytogenetic risk groups.

### TGIF1 expression correlated with survival in independent cohorts of AML patients

To substantiate the association between *TGIF1* expression and survival observed in the discovery group, we reanalyzed hybridization data from a published study *^23^* and related leukemia cell *TGIF1* expression to survival in this larger (n = 285) group of patients. When patients were segregated into 5 groups according to myeloblast *TGIF1* RNA abundance and trend and log-rank analysis of survival in these patients was carried out, a concentration- or dose-dependence in survival was observed (Figure 2B). We also investigated whether *TGIF1* expression differed in five patient clusters, 5, 9, 10, 12, and 13 *^23^*. Cluster 5 consisted of FAB class M4 or M5 leukemias, cluster 10 expressed the *EVI1* oncogene, and clusters 9, 12 and 13 were defined by the presence of (inv)16, t(15;17), and t(8;21), respectively. Consistent with the association of t(15;17), t(8;21), and inv(16) with a better outcome and the inverse association between *EVI1* expression and survival *^14^*, OS was shorter in patients in clusters 5 and 10 compared to 9, 12, and 13 *^23^* (Figure 2B). Mean *TGIF1* levels differed significantly between these clusters (P < 0.0001), with significant inter-group differences noted between clusters 5 and 9 (P = 0.0001), 5 and 12 (P < 0.0001), 5 and 13 (P < 0.0001), 10 and 12 (P < 0.0001), and 10 and 13 (P = 0.0003).

Finally, OS was significantly worse in AML patients from The Cancer Genome Atlas (TCGA) whose *TGIF1* expression was below the median than those with expression above the median for both probe set 1566901_at (P < 0.0001) and 203313_s_at (P = 0.000254) (http://servers.binf.ku.dk/bloodspot/?gene=TGIF1&dataset=normal_human_v2_with_AMLs) (Supplemental Figure S1). This association was specific for *TGIF1*, as there was no difference in survival according to expression of the related *TGIF2* gene. Thus, analysis of multiple, independent groups of patients confirmed the correlation between *TGIF1* gene expression and outcome and, for the European cohort, the quantitative nature of this relationship.

### TGIF1 expression was unrelated to the presence of FLT3 gene mutations

To determine whether decreased *TGIF1* expression was related to *FLT3* mutation, a widely used prognostic factor in adults with AML, RNA from patient cells and the MV4-11 cell line, used as a positive control for the *FLT3* ITD *^24^*, were characterized by RT-PCR analysis. Of the 55 patients from whom material was available for analysis, a *FLT3* ITD was detected in 15 (27%) and the D835Y missense mutation in two (4%). In two patients with a *FLT3* ITD, no wild-type *FLT3* RNA could be detected at the sensitivity of this method, and in accord with published data *^25^*, these individuals had very short survivals. In contrast, the OS of patients expressing mutant and wild-type *FLT3* mRNA did not differ significantly from patients expressing only wild-type message in our analysis (not shown). Importantly, no association between level of *TGIF1* expression and *FLT3* mutation could be discerned (data not shown). These data thus argue against a model in which mutational activation of FLT3 signaling represses *TGIF1* expression.

### Reduced TGIF1 expression in myeloid leukemia cells was not associated with sequence variants

The association between decreased *TGIF1* expression and shorter survival could be compatible with a tumor suppressor function for this transcription factor. Since well-characterized loss-of-function mutations in this gene have been described in a human genetic disorder, we undertook analysis of *TGIF1* coding and 5’ untranslated sequence for these or other mutations in 49 patients in the discovery group. Although non-synonymous as well as synonymous sequence variants were detected in 18 patients (37%), with previously unreported single nucleotide polymorphisms found in five (Supplemental Table S2), none of these corresponded to the mutations observed in patients with holoprosencephaly. Likewise, Southern blot analysis did not reveal *TGIF1* gene rearrangements in any patient studied (data not shown). Thus, no definite abnormality in either gene sequence or structure was detected in any individual with AML.

When *TGIF1* mRNA abundance in myeloblasts was examined in aggregate, expression appeared to vary in continuous fashion (Supplemental Figure S2). *TGIF1* expression in 100 lymphoblastoid cell lines derived from normal individuals likewise appeared to be a continuous variable, with an approximately 200-fold difference between the lowest and highest expresser (Supplemental Figure S3).

### Tgif1 gene loss decreased survival in an experimental model of acute myeloid leukemia

Given our finding that *TGIF1* expression correlated significantly with survival in AML, we sought to determine if myeloblast *TGIF1* abundance affected any aspect of leukemia cell biology. Our previous study has shown that *Tgif1* inactivity promotes the quiescence and the self-renewal activity of hematopoietic stem cells (HSC) without disturbing steady-state hematopoiesis using a *Tgif1* knockout model*^26^*. To understand the role of *Tgif1* in leukemia cells, in the present study, AML was induced by introduction of an *MLL-AF9* fusion cDNA. Specifically, lineage-negative bone marrow cells (BMCs) from *Tgif1*^+/+^ and *Tgif1*^-/-^ mice were transduced with the MLL-AF9- GFP retrovirus and transduced cells transplanted into sub-lethally irradiated recipients *via* tail vein injection. Development and progression of leukemia in recipient mice was then monitored by flow cytometry analysis of green fluorescent protein (GFP)-expressing nucleated cells in peripheral blood. Six weeks post-transplant, mice transplanted with *Tgif1*^-/-^ BMCs showed higher numbers of GFP^+^ myeloid (CD11b^+^ and Gr-1^+^) cells (Figure 3A and 3B) but lower numbers of B cells (Figure 3C) compared to mice transplanted with *Tgif1*^+/+^ BMCs. Necropsy showed that all mice with circulating donor-derived leukemia cells had splenomegaly, with massive infiltration of myeloblasts observed in both spleen and bone marrow (data not shown).

Importantly, Kaplan-Meier analysis showed that mice transplanted with MLL-AF9- transduced *Tgif1*^-/-^ BMCs survived a significantly shorter time than mice transplanted with transduced *Tgif1*^+/+^ BMCs (Figure 4). As mice transplanted with either *Tgif1*^+/+^ or *Tgif1*^-/-^ BMCs succumbed to leukemia with very short latency, the aggressive nature of this leukemia almost certainly underestimated the potential effect of *Tgif1* gene loss on survival and LSC number (see below). Regardless, these results are consistent with the patient data (Figures 1 and 2) and indicate that *Tgif1* abundance impacts progression of MLL-AF9-induced AML.

**Figure 3.**
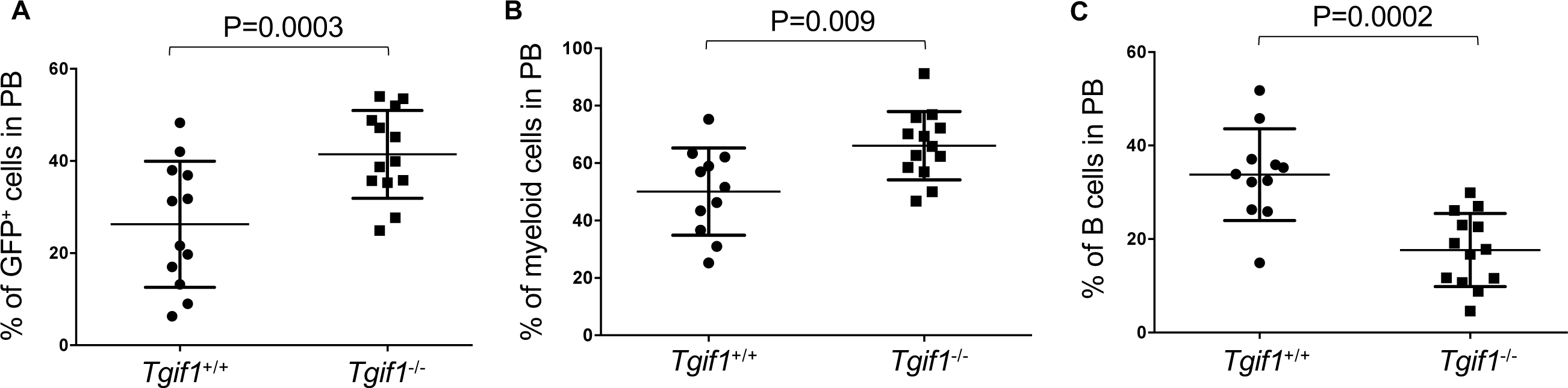
MLL-AF9-induced AML in *Tgif1*^-/-^ mice was more aggressive than in *Tgif1*^*+/+*^ mice. Lin^-^ bone marrow cells from *Tgif1*^+/+^ *or Tgif1*^-/-^ mice were transduced with MLL-AF9-GFP retrovirus and transduced cells transplanted into sub-lethally irradiated *Tgif1*^+/+^ and *Tgif1*^-/-^ recipient mice, respectively. Six weeks post-transplant, the percentages of **A**, GFP^+^ cells; **B**, myeloid cells (CD11b^+^ and Gr-1^+^) and **C**, B-lymphocytes (B220^+^) in peripheral blood (PB) were analyzed by flow cytometry.

**Figure 4.**
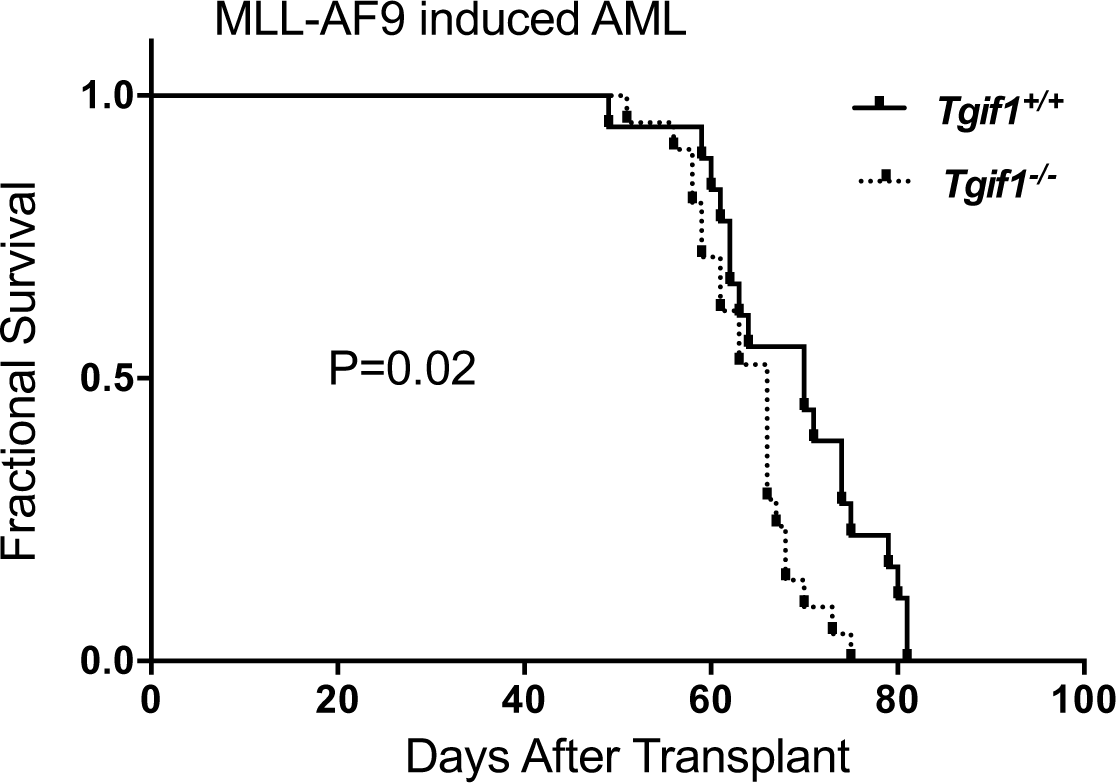
*Tgif1* loss was associated with shorter survival *in vivo*. Kaplan-Meier survival curves for mice with the indicated genotype and MLL-AF9-induced AML. Lin^-^ bone marrow cells from *Tgif1*^+/+^ and *Tgif1*^-/-^ mice were transduced with MLL-AF9- GFP retrovirus and transduced cells transplanted into sub-lethally irradiated *Tgif1*^+/+^ and *Tgif1*^-/-^ recipients, respectively.

### Tgif1 gene loss increased leukemia initiating cell frequency in MLL-AF9-induced AML

LICs are thought to be crucial for the maintenance of AML *in vivo ^27-29^*. Therefore, one explanation for the shorter survival seen with *Tgif1* loss could be higher numbers of leukemia initiating cells. To investigate, we carried out limiting dilution analysis of LIC number, transplanting serial numbers of MLL-AF9-transduced *Tgif1*^+/+^ or *Tgif1*^-/-^ spleen cells from primary recipients into non-conditioned C57BL/6J mice. Indeed, there was a two-fold higher frequency of LICs in *Tgif1*^-/-^ mice with AML (1 in 125) (95% CI: lower; 1 in 279 and higher; 1 in 56.5) than in *Tgif1*^+/+^ mice (1 in 250) (95% CI: lower; 1 in 603 and higher; 1 in 103.6). These data suggest that *Tgif1*^-/-^ leukemic populations were enriched in LIC function and/or number. Consistent with this notion, mice transplanted with *Tgif1*^-/-^ leukemia cells over a range of doses had inferior survival compared to mice transplanted with *Tgif1*^+/+^ cells (Figure 5 A-D).

### Tgif1 loss resulted in earlier relapse and more aggressive disease

Since leukemia cell preparations from *Tgif1*^-/-^ mice contained higher frequencies of LICs, we predicted that disease induced by these cells would relapse earlier and progress more rapidly after treatment than disease in mice receiving *Tgif1*^+/+^ leukemia cells. To simulate the clinical setting, we transplanted *Tgif1*^+/+^ and *Tgif1*^-/-^ MLL-AF9-transformed-spleen cells from mice with established leukemia into sub-lethally irradiated recipients and treated these mice with cytarabine and doxorubicin chemotherapy as described *^30^*. For mice transplanted with *Tgif1*^-/-^ leukemia cells, GFP-expressing cells appeared earlier and in higher numbers in peripheral blood than in mice transplanted with *Tgif1*^+/+^ leukemia cells (Figure 6A). While treatment extended survival of both *Tgif1*^+/+^ and *Tgif1*^-/-^ mice with AML (compare Figure 6B to Figure 5D), mice transplanted with *Tgif1*^-/-^ leukemia cells still had shorter survival than mice transplanted with *Tgif1*^+/+^ leukemia cells (Figure 6B). Although selection at the level of a stem cell is not proven by these data, the differences in latency of disease and response to chemotherapy, we posit, are due to the higher frequency and/or greater fitness of LICs and more rapid expansion of leukemia in *Tgif1*^-/-^ mice (see Figure 6A).

**Figure 5.**
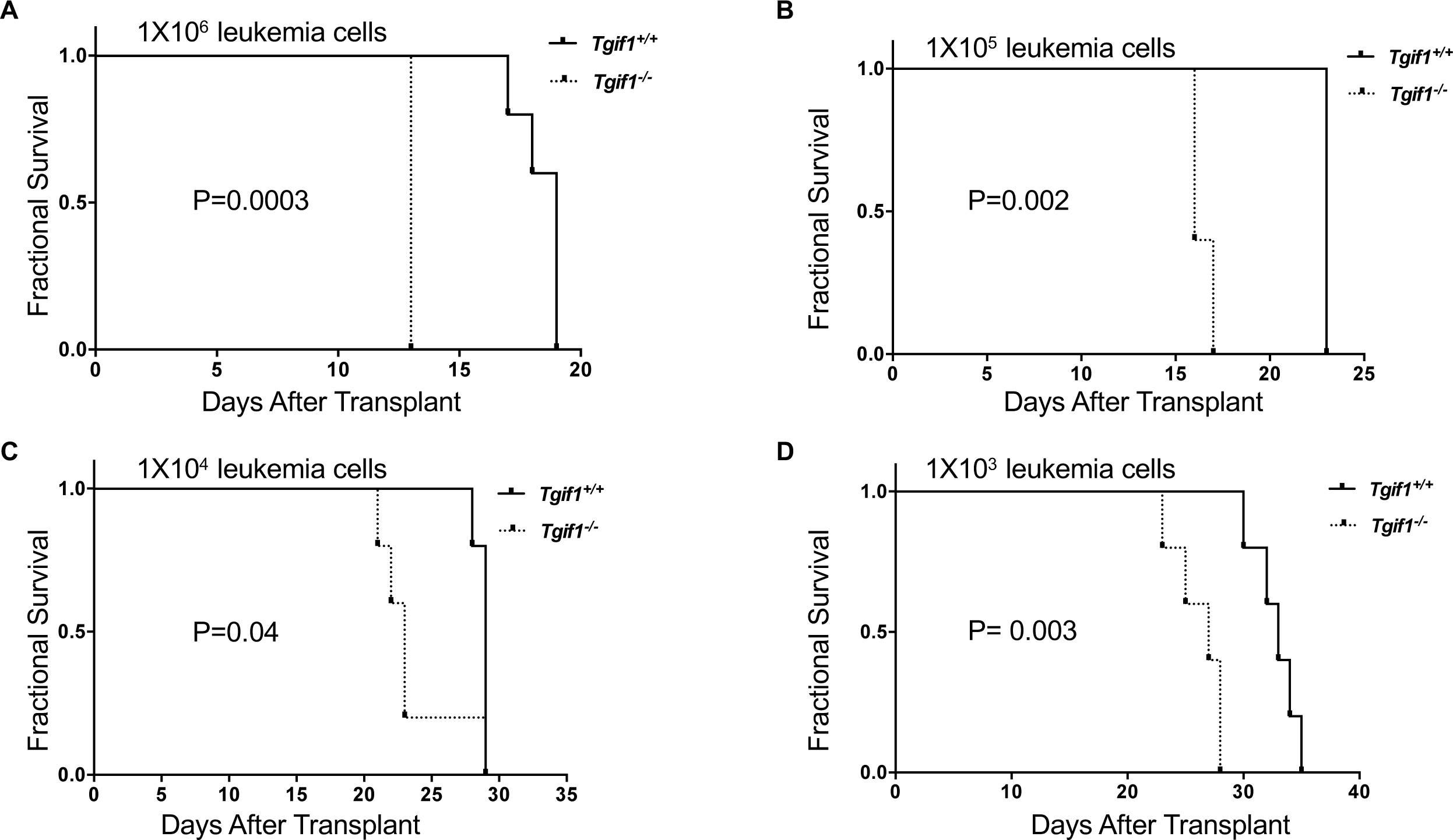
LSC frequency was increased in*Tgif1*^-/-^ mice with AML compared to *Tgif1*^+/+^ mice. Lin^-^ bone marrow cells from *Tgif1*^+/+^ and *Tgif1*^-/-^ mice were transduced with MLL-AF9-GFP retrovirus and transduced cells transplanted into sub-lethally irradiated *Tgif1*^+/+^ and *Tgif1*^-/-^ recipients, respectively. At a predetermined time, serial dilutions of spleen cells from mice with leukemia then collected and transplanted into secondary recipients. Kaplan-Meier analysis of survival is plotted as a function of number of transplanted spleen cells. **A**, 1×10^6^; **B**, 1×10^5^; **C**, 1×10^4^; **D**, 1×10^3^.

**Figure 6.**
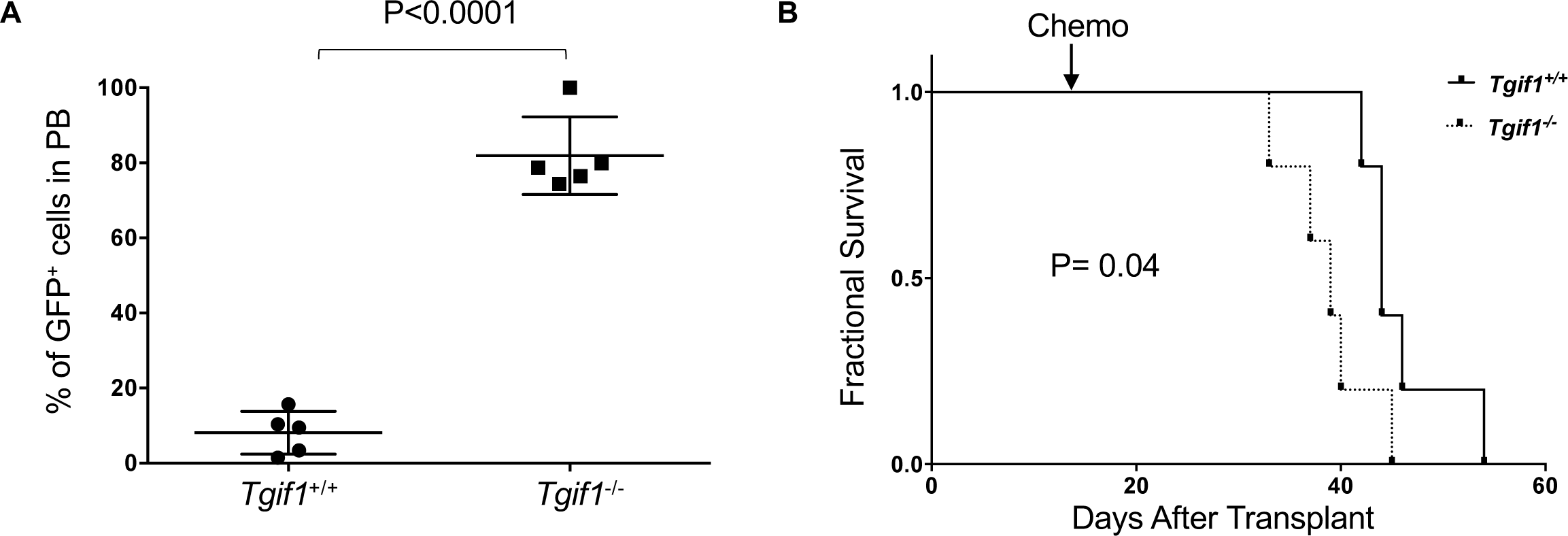
AML in *Tgif1*^-/-^ mice progressed earlier after chemotherapy than in *Tgif1*^*+/+*^ AML mice. Sub-lethally irradiated C57BL/6 mice were transplanted with spleen cells from *Tgif1*^+/+^ or *Tgif1*^-/-^ mice with established MLL-AF9-induced AML, and two weeks following transplant, recipient mice were treated with doxorubicin for 3 days and cytarabine for 5 days by intraperitoneal injection. **A**, Percentage of GFP^+^ cells in peripheral blood two weeks post chemotherapy. **B**, Kaplan-Meier analysis of survival of *Tgif1*^+/+^ and *Tgif1*^-/-^ mice with AML after chemotherapy.

### Tgif1 loss decreased survival in a model of chronic myeloid leukemia

Given data suggesting *Tgif1* expression impacted survival in AML, we went on to investigate whether *Tgif1* loss also affected leukemia cell biology in a mouse model of CML, a disease for which there is arguably the greatest evidence for a malignant stem cell. To that end, lineage marker-negative (Lin^-^) BMCs from *Tgif1*^+/+^ and *Tgif1*^-/-^ mice were transduced with the BCR-ABL-GFP retrovirus, which were transplanted into sub- lethally or lethally irradiated recipient mice. After transplant, leukemia progression was monitored from the kinetics of GFP-expressing myeloid cells in peripheral blood (PB) of recipient mice. Six weeks post-transplant, *Tgif1*^-/-^ mice showed higher numbers of GFP^+^ cells in PB and BM, including the LSK population, compared to *Tgif1*^+/+^ mice (Supplemental Figure S4 A, B & C, respectively). Kaplan-Meier analysis revealed that mice transplanted with BCR-ABL-GFP-transduced *Tgif1*^-/-^ BMCs showed significantly shorter survival than mice transplanted with BCR-ABL-GFP transduced *Tgif1*^+/+^ BMCs (Figure 7A). When recipient mice were conditioned with a lower dose of irradiation, leukemia developed with a longer latency; however, *Tgif1* genotype still significantly impacted survival (Figure 7B). Interestingly, mice transplanted with BCR-ABL-transduced haploinsufficient (*Tgif1^+/-^*) BMCs also had shorter survivals (Supplemental Figure S5), consistent with the greater long-term repopulating ability of *Tgif1*^+/-^ HSCs compared to *Tgif1*^+/+^ HSCs after transplantation*^26^*. In sum, these data are compatible with the patient and mouse AML data and suggest *Tgif1* expression could also impact survival in CML.

**Figure 7.**
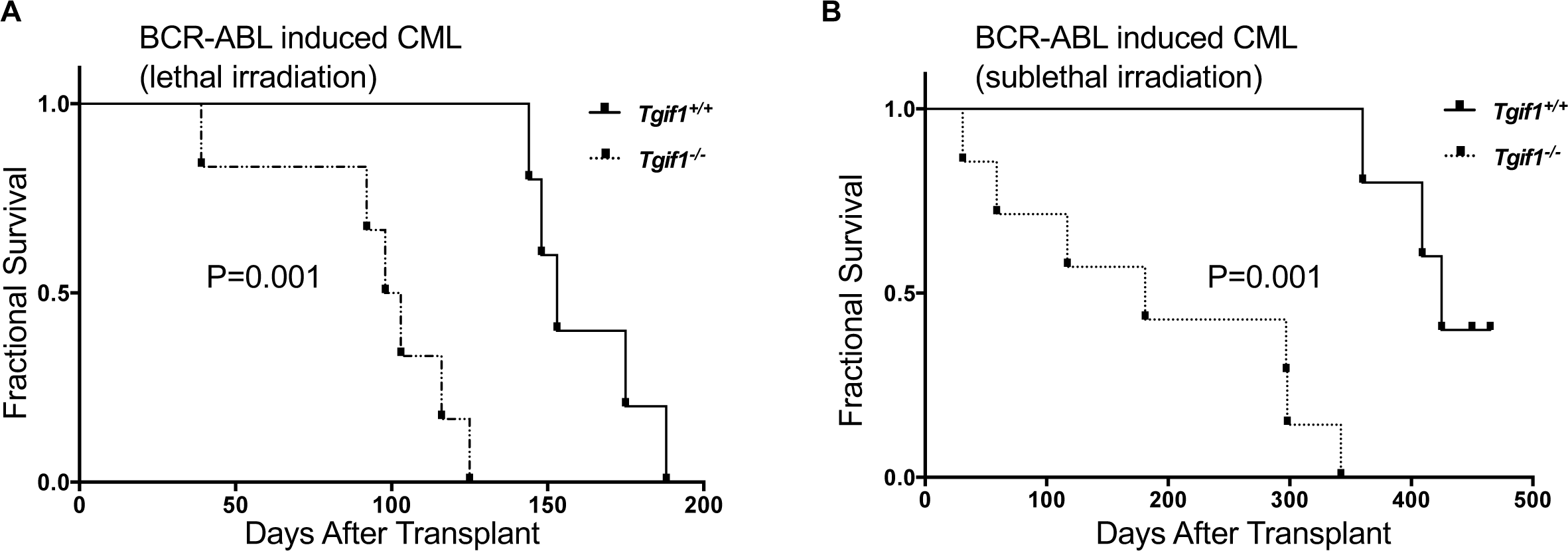
*Tgif1*^-/-^ mice with BCR-ABL-induced CML had inferior survival compared to *Tgif1*^*+/+*^ mice with CML. Lin^-^ c-Kit^+^ bone marrow cells from *Tgif1*^+/+^ or *Tgif1*^-/-^ mice were transduced with BCR-ABL-GFP retrovirus and transduced cells transplanted into C57BL/ 6J recipient mice. Recipient mice were **A**, lethally irradiated; **B**, sub-lethally irradiated.

### Tgif1 gene loss affected multiple transcriptional networks in AML

To gain insight into the biological and molecular pathways affected by *Tgif1* gene loss, we compared global gene expression profiles in *Tgif1*^-/-^ and *Tgif1*^+/+^ myeloid leukemia cells using RNA-seq analysis. We identified at least 45 genes that were differentially expressed in these two populations – 33 of these were significantly upregulated and 12 were down-regulated in *Tgif1*^-/-^ relative to *Tgif1*^+/+^ leukemia cells (Table 1).

**Table 1.** Differentially expressed genes in *TGIF1*^-/-^vs. *TGIF1*^+/+^ leukemic cells

| ID | Symbol | Location | Log Ratio |
| --- | --- | --- | --- |
| **Slc15a2** | SLC15A2 | Plasma Membrane | -7.651 |
| **Eps8l1** | EPS8L1 | Cytoplasm | -7.587 |
| **Dlgap1** | DLGAP1 | Plasma Membrane | -3.719 |
| **Gdf3** | GDF3 | Extracellular Space | -3.658 |
| **Camk2b** | CAMK2B | Cytoplasm | -3.619 |
| **Col19a1** | COL19A1 | Extracellular Space | -3.488 |
| **Kcnq5** | KCNQ5 | Plasma Membrane | -2.727 |
| **Vill** | VILL | Cytoplasm | -2.499 |
| **Cgnl1** | CGNL1 | Plasma Membrane | -1.938 |
| **Cdkn2c** | CDKN2C | Nucleus | -1.649 |
| **Gm15448** | LILRB3 | Plasma Membrane | -1.362 |
| **Selm** | SELM | Cytoplasm | -0.761 |
| **Ly75** | LY75 | Plasma Membrane | 0.652 |
| **Anxa3** | ANXA3 | Cytoplasm | 0.714 |
| **Agap1** | AGAP1 | Cytoplasm | 0.742 |
| **Ttc21a** | TTC21A | Extracellular Space | 0.967 |
| **Pip5k1b** | PIP5K1B | Cytoplasm | 0.976 |
| **B4galt6** | B4GALT6 | Cytoplasm | 1.029 |
| **Ptgs1** | PTGS1 | Cytoplasm | 1.045 |
| **Olfm4** | OLFM4 | Extracellular Space | 1.123 |
| **Dsp** | DSP | Plasma Membrane | 1.212 |
| **Gfi1** | GFI1 | Nucleus | 1.278 |
| **Plekha6** | Plekha6 | Other | 1.291 |
| **Sept5** | SEPT5 | Cytoplasm | 1.301 |
| **Optn** | OPTN | Cytoplasm | 1.325 |
| **Kcnh7** | KCNH7 | Plasma Membrane | 1.417 |
| **Fcnb** | FCN1 | Extracellular Space | 1.427 |
| **S100a8** | S100A8 | Cytoplasm | 1.503 |
| **Gca** | GCA | Cytoplasm | 1.575 |
| **Serpine2** | SERPINE2 | Extracellular Space | 1.875 |
| **Camp** | CAMP | Cytoplasm | 1.940 |
| **Serpine1** | SERPINE1 | Extracellular Space | 2.160 |
| **Retnlg** | Retnlg | Extracellular Space | 2.167 |
| **Cd74** | CD74 | Plasma Membrane | 2.609 |
| **C1qb** | C1QB | Extracellular Space | 3.161 |
| **C1qc** | C1QC | Extracellular Space | 3.269 |
| **Adam12** | ADAM12 | Plasma Membrane | 3.167 |
| **C1qa** | C1QA | Extracellular Space | 3.503 |
| **Mmp14** | MMP14 | Extracellular Space | 4.032 |
| **Thbs4** | THBS4 | Extracellular Space | 4.578 |
| **Bmp1** | BMP1 | Extracellular Space | 4.577 |
| **Npr1** | NPR1 | Plasma Membrane | 5.607 |
| **Mtus2** | MTUS2 | Other | 5.713 |
| **Syce1** | SYCE1 | Nucleus | 7.905 |
| **4930447C04Rik** | C14orf39 | Extracellular Space | 8.430 |

Ingenuity Pathway Analysis (IPA) allowed us to interrogate which upstream regulators contributed to gene expression changes. In this analysis, genes involved in TGF-β signaling were significantly enriched (Figure 8). Other regulators identified by this analysis included all-*trans* retinoic acid (ATRA) (P = 1.02 × 10^−8^) and serum response factor (SRF) (P = 5.34 × 10^−6^). Like the TGF-β targets, genes activated by ATRA were upregulated by *Tgif1* loss, consistent with this transcription factor’s action as a corepressor of retinoic acid receptor (RAR)-dependent transcription.

**Figure 8.**
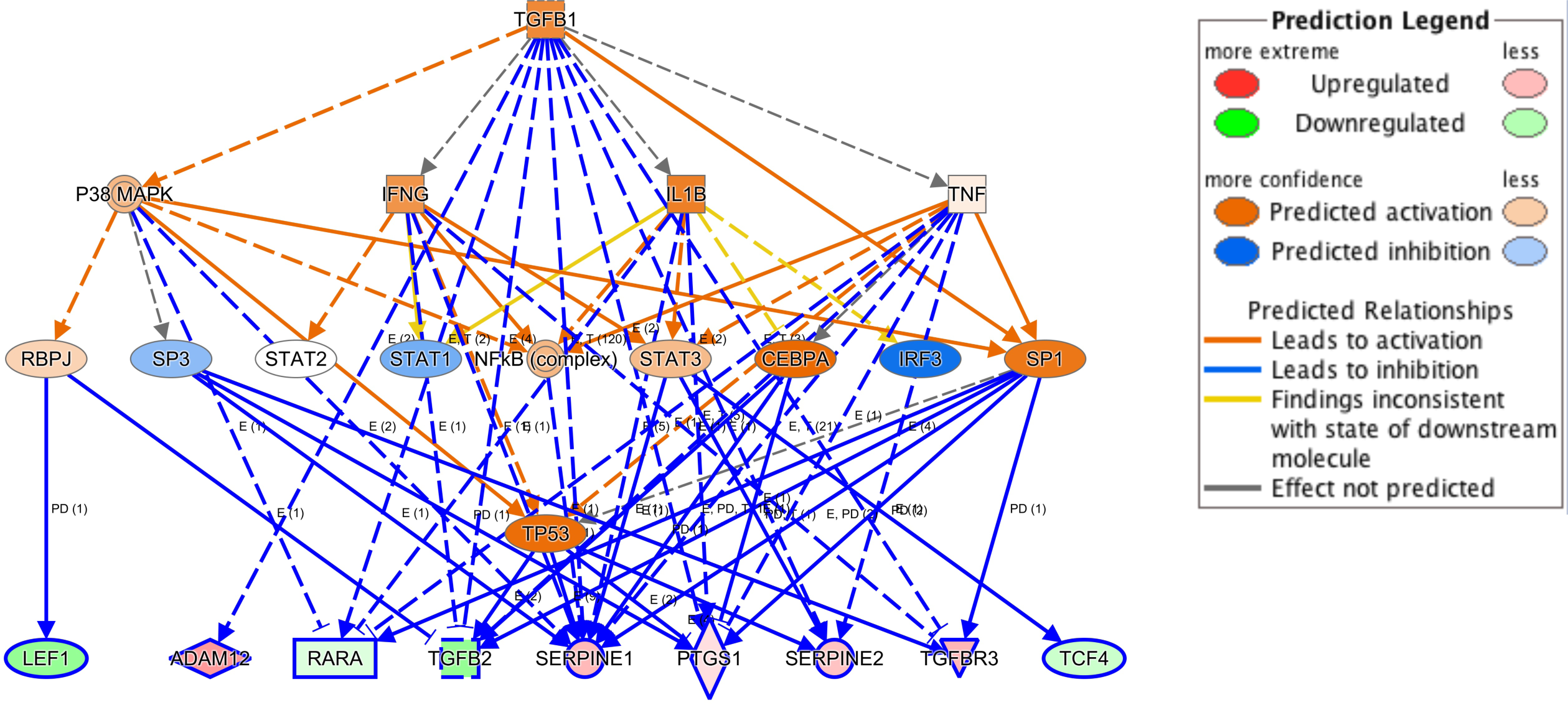
Upstream regulator analysis of differentially expressed genes. This analysis identified TGF-b as a major regulator of differentially expressed genes (p=8.52 × 10^−10^).

Genes important for embryonic stem cell pluripotency were highlighted in IPA canonical pathway analysis (P = 5.88 × 10^−4^) (Supplemental Figure S6), while hematological system development and function (P = 1.05 × 10^−7^) was enriched in IPA physiological functions, unsurprising given the cellular source of these RNAs. Other functions impacted by *Tgif1* expression included cellular movement (P = 1.99 × 10^−9^), leukocyte function (P = 3.87 × 10^−8^), cell death and survival (P = 5.38 × 10^−8^), myeloid cell function (P = 3.24 × 10^−8^), and cancer (P = 2.63 × 10^−7^). In summary, pathway analysis in *Tgif1* knockout leukemia cells revealed upregulation of TGF-β signaling, with potentially important consequences for LSC function.

## Discussion

Our studies revealed that expression of the transcriptional regulator *TGIF1* correlates quantitatively with survival in adults with AML treated with standard induction and consolidation chemotherapy, a finding that was validated in two other, independent cohorts of patients. *TGIF1* expression impacted outcome in each cytogenetic risk group, and, at least for patients treated at our institution, was an independent risk factor in multivariate analysis.

Consistent with these results, we found that loss or haploinsufficiency of this gene accelerated leukemia development, impaired response to chemotherapy, and reduced survival in mouse models of acute and chronic myeloid leukemia. Whether quantification of *TGIF1* expression will have clinical utility in risk stratification in patients with AML, however, requires further investigation.

TGIF1 belongs to the TALE family of homeodomain proteins *^21^* and functions as a corepressor of retinoic acid and TGF-β-stimulated transcription. It does so by interfering with retinoid X receptor (RXR) binding to DNA and recruiting general corepressors *^31^* and promoting recruitment of histone deacetylases to the TGF-β signaling intermediate Smad2 *^20-22, 32^*, respectively. TGIF1 can also inhibit transcription directly through binding to a TG-rich sequence element *via* its homeobox domain. Inactivating mutations in *TGIF1* cause autosomal dominant holoprosencephaly *^33-36^*, the most common inherited defect in forebrain development in humans, while inactivation of the *Tgif1* gene in mice, complete or partial, increases quiescence in bone marrow HSCs and enhances long-term repopulating activity without an effect on steady-state hematopoiesis *^26^*.

In light of studies demonstrating that LSC frequency at diagnosis in AML correlates with increased minimal residual disease and poor survival *^37^*, the finding that reduced or absent expression of Tgif1 in mouse models of myeloid leukemia enhanced LSC function provides at least one explanation for how it affects recurrence. Since effective therapy after relapse is lacking in AML, except for hematopoietic stem cell transplantation in the case when a second remission can be induced, this explains how patient survival would be impacted.

We found using a mouse model of AML that loss of *Tgif1* resulted in a shorter latency in development of leukemia, earlier relapse after chemotherapy, and inferior survival. In addition, we found in a model of CML that heterozygous as well as homozygous loss of *Tgif1* shortened survival. These results underscore the quantitative relationship between *TGIF1* abundance and survival in AML patients and stem cell frequency in mice. They are also compatible with a recent report demonstrating that enforced expression of *TGIF1* decreased leukemia cell proliferation, induced terminal differentiation, and increased survival in MLL-AF9 leukemia *^38^*. Altogether, these data establish a biological basis for the association between *TGIF1* expression and clinical outcome.

In accord with its function as transcriptional repressor or corepressor, RNA expression profiling identified a number of genes that were upregulated in *Tgif1-*null leukemia cells, with almost half the genes differentially expressed in knockout and wild-type leukemia cells involved directly or indirectly in TGF-β signaling (Supplemental Figure 6). This result also corroborates the enhanced TGF-β signaling and/or target gene expression noted in epithelial *^32^* and myeloid leukemia cells *^39^* in which TGIF1 expression was reduced by RNA interference. Importantly, expression of the prototypic TGF-β target gene, *plasminogen activator inhibitor-1*, in human myeloid leukemia cells correlated significantly with expression of a type I TGF-β receptor, activin receptor-like kinase-5 (ALK-5), and presumably downstream signaling strength. Likely for the reason offered here, *ALK-5* expression, in turn, correlated inversely with achievement of CR and with EFS, RFS, and OS *^40^*.

Relevant to this work, TGF-β was shown to promote self-renewal in human HSCs *^41^* and quiescence in murine HSCs *^42^* and to enhance the function of leukemia stem cells *^41, 43-55^*, which could also explain the curative potential of a small-molecule inhibitor of the type I TGF-β receptor *^56, 57^* added to a BCR-ABL tyrosine kinase inhibitor in CML in mice *^51^*. Together with the gene expression results, these studies predict, therefore, that a TGF-β signaling inhibitor, most logically used in combination with post-remission chemotherapy, would have the greatest impact in patients with the lowest *TGIF1* expression.

In addition to an effect on TGF-β target gene expression, loss of TGIF1 caused apparent derepression of retinoic acid receptor (RAR) target genes, consistent with its ability to act as a corepressor of RAR/RXR-mediated transcription. Finally, our results do not exclude direct effects of TGIF1 on gene expression or indirect effects through interaction with other TALE homeodomain proteins, MEIS1 for example *^38^*. As chemical inhibitors are available for TGF-β or RAR signaling, it should be possible to determine which of these signaling pathways is most important for TGIF1’s actions in leukemia cells. It will also be important to determine whether inhibition of these pathways, and TGF-β in particular, would have equivalent biological effects on leukemic compared to normal stem cells, which would have therapeutic implications.

Although loss-of-function mutations in *TGIF1* have been recognized in holoprosencehaly *^35,36^*, we did not observe any coding or promoter mutations or any structural rearrangements of the gene in AML patients. Furthermore, individuals with holoprosencephaly and *Tgif1*-null mice are not at increased risk for development of myeloid leukemia, all of which argues against a function of this protein as a tumor suppressor. Its graded expression in hematopoietic cells and dose-dependent effect on cell function, most clearly demonstrated for normal HSCs *^26^*, suggest that *TGIF1* acts as a quantitative trait locus in normal and malignant stem cell biology. In addition, *TGIF1* exemplifies a type of cancer modifier gene that affects disease progression and persistence rather than pathogenesis.

In summary, by relating transcriptional profiles in myeloblasts from patients with AML to survival, we have identified a new prognostic marker relevant, potentially, to all myeloid leukemias. In addition, *TGIF1* may be the prototype of a stem cell modifier gene, operating not in the initiation of leukemia but in disease progression and persistence.

## Materials and Methods

### Patients

Pre-treatment samples from 60 patients with AML treated at Vanderbilt University Hospital or Nashville VA Medical Center from 1996-2000 were obtained with the informed consent of patients and approval of the responsible Institutional Review Board (Supplemental Table 1) *^47^*. A diagnosis of AML was made by examination of blood and/or bone marrow with cytochemical stains and with cytogenetics and flow cytometry analysis. Mononuclear cells were enriched by Ficoll-Hypaque centrifugation, depleted of T-cells and monocytes with antibodies to CD2 and CD14 (Dynal Biotech, Brown Deer, WI), respectively, and stored frozen at −80°C. Anti-CD2-conjugated beads alone were used for leukemias of FAB classes M4 and M5.

### Classification by cytogenetic risk group

Patients were risk-stratified according to the results of cytogenetics analysis. The good-risk group was defined by the presence of t(8;21), t(15;17), or inv(16) and poor-risk group by del(5), del(7), t(6,9), alterations in 11q23, or multiple cytogenetic abnormalities. Patients with normal karyotypes were assigned to the intermediate-risk group.

### Survival analysis in patients

Gene expression profiles were related to individual patient survival. Modified Cox PH regression analysis was used to rank genes by ability to predict OS, with the importance of individual genes weighted in accordance with the data available. Survival analysis in the discovery group was carried out using SPSS version 22 (Chicago, IL). Patients were enrolled between 1996 and 2001 and their outcomes last updated on June 6, 2004. OS was calculated from the time of initial diagnosis (n = 53) or relapse (n = 7) to last follow-up or death. Survival was analyzed as a function of *TGIF1* expression using both normalized hybridization and real-time PCR data. For the original microarray analysis, *TGIF1* expression in leukemia cells was divided by that in a commercial reference, log_2_ transformed, and related to patient survival. Kaplan-Meier plots were analyzed with a log-rank test. P values were based on two-sided comparisons, with P < 0.05 considered significant. RFS was calculated from the time of diagnosis to documented relapse, SCT, death, or last follow-up. Patients who failed to achieve a CR were considered to have an RFS of 0 and all untreated patients were excluded. As described below, the patient cohort described in the study by Valk *et al ^23^* was subdivided into five groups according to leukemia cell *TGIF1* expression and the actuarial probabilities of OS and event-free survival (EFS) were estimated from Kaplan-Meier analysis (Stata Statistical Software, release 9.2; College Station, TX). Trend and log-rank analysis were carried out using Microsoft Excel (Redmond, WA). Hybridization data were normalized with MAS 5.0, log_2_-transformed *TGIF1* expression ratios were median-centered, and differences in *TGIF1* expression between clusters 5, 9, 10, 12, and 15 were assessed by one-way analysis of variance (SAS, Cary, NC).

### Treatment

Ninety four percent (50/53) of newly diagnosed patients received induction chemotherapy, with 82% (41/50) treated with 100 mg/m^2^/day of cytarabine by continuous infusion from days 1-7 and 12 mg/m^2^ of idarubicin or 45 mg/m^2^ daunorubicin intravenously on days 1-3. Two other patients received cytarabine for five days instead of seven. Most of the remaining patients received cytarabine and an anthracycline together with other agents. Five patients received an induction regimen of dexamethasone, cytarabine, thioguanine, etoposide, and daunorubicin. Four patients with *de novo* disease did not receive induction chemotherapy, while three of seven patients with relapsed leukemia received salvage therapy.

### Quantitative real-time PCR analysis

Total cellular RNA (1 µg) was transcribed into cDNA with a mixture of random hexamer and oligo(dT) primers (iScript cDNA Synthesis Kit. Bio-Rad, Hercules, CA). Real-time PCR analysis was carried out using SYBR Green I detection (iTaq SYBR Green Supermix and MyiQ Single-Color Real-Time PCR Detection System, Bio-Rad).

Amplification parameters consisted of an initial denaturation at 95°C for 3 min. This was followed by 35 cycles of denaturation at 94°C for 30 seconds, annealing at 56.7°C for 30 seconds, and extension at 75°C for 10 seconds. *TGIF* expression in leukemia samples was quantified with the same reference RNA employed for the microarray studies, and relative quantification of *glyceraldehyde phosphate dehydrogenase* (*GAPDH*) gene expression was performed identically. Threshold cycles (C_T_) were determined for *GAPDH* and *TGIF*, and if the *GAPDH* C_T_ differed by three cycles or more between blast cell and reference cDNA, the concentration and quality of RNA in that patient’s sample was reevaluated. Four of the five samples to which this applied were subsequently used after repeat determination of RNA concentration. Primers were designed using the MacVector software package (Accelrys, San Diego, CA). The sequences of the primers used in this analysis were: forward *TGIF* primer CAGATTCTTCGGGATTGGCTG, reverse *TGIF* primer GGCGTTGATGAACCAGTTACAGAC, forward *GAPDH* primer AATCCCCAGTGCTCCTTTTC, reverse *GAPDH* primer CAAAGTTGTCATCGATGACC.

### TGIF mutational analysis

cDNA synthesis was carried out with the Superscript First-Strand System (Invitrogen Life Technologies, Carlsbad, CA) using 1 µg of total cellular RNA and an oligo(dT) primer. One-tenth volume of the first-strand reaction was then used as template for PCR amplification (Elongase Amplification System, Invitrogen). The sequences of the primers used in this analysis, corresponding to nucleotides 25-45 and 1489-1509, respectively, of *TGIF* RNA sequence, were: forward primer GCAGGAGCAGGGAACAAAG, reverse primer CAGTATGTGGGCATCCTGTTC. Amplification parameters consisted of an initial denaturation at 94°C for 3 min, annealing at 56°C for 30 seconds, and extension at 72°C for 1 minute. This was followed by 35 cycles of denaturation at 94°C for 30 seconds, annealing at 56°C for 30 seconds, and extension at 68°C for 90 seconds and, terminally, extension at 68°C for 5 minutes. The resulting 1,477 bp cDNA product was purified in a Microcon 50 microconcentrator (Amicon, Danvers, MA) and an aliquot visualized by ethidium bromide staining after electrophoresis in a 1% agarose gel. Automated sequencing of purified PCR products was carried out with a BigDye Terminator cycle sequencing kit (v3.1, Applied Biosystems) using sets of primer encompassing the complete *TGIF* coding sequence. All sequence variants were confirmed by sequencing of both strands.

### FLT3 mutational analysis

The presence of an ITD or D835Y kinase domain mutation in *FLT3* was determined by reverse transcription-polymerase chain (RT-PCR) analysis of leukemic cell RNA. Total cellular RNA was purified using the RNeasy purification system (Qiagen) from blasts obtained as described above. RNA was reverse transcribed using an iScript cDNA Synthesis kit according to the manufacturer’s (Bio-Rad) instructions, and one-tenth volume of the RT mixture was used for PCR amplification. The sequences of the primers used in this analysis were: forward ITD TGGTGTTTGTCTCCTCTTCATTGTCG, reverse ITD TCTTTCAGCATTTTGACGGCAACC, forward D835Y GAAGAAGAGGAGGACTTGAATGTGC, reverse D835Y AGGTCGCCTGTTTTGGTAGGTG. PCR was carried out with TaKaRa LA Taq polymerase (TaKaRa Mirus Bio, Madison, WI) in a 50 μl reaction volume containing 1× PCR buffer, 2mM MgCl_2_, 0.4 mM of each dNTP, 0.2 μM of each primer, and 2.5 U of enzyme. Amplification parameters consisted of an initial denaturation at 94°C for 3 min, annealing at 56°C for 30 seconds, and extension at 72°C for 1 minute, followed by 30 cycles of denaturation at 94°C for 30 seconds, annealing at 56°C for 30 seconds, and extension at 72°C for 1 minute. PCR products were digested with *Eco*RV for 1 hour at 37°C for detection of the D835Y mutation. Amplified fragments and *Eco*RV-digested products were analyzed on a 3% NuSieve-agarose gel (Cambrex, Rutherford, NJ), with a *FLT3* ITD recognized from an increase in the size of the amplified fragment (>300 bp) and the D835Y mutation by the absence of *Eco*RV digestion.

### Evaluation of remission

Bone marrow aspiration and biopsy were carried out on days 14 and 28 of therapy to assess response. CR was defined as the disappearance of leukemia cells from the bone marrow accompanied by return of normal blood counts. For patients with antecedent MDS, CR was defined as recovery of pre-leukemic numbers, with blasts making up ≤ 5% of nucleated bone marrow cells.

### Mice and plasmids

Mice with a *Tgif1* null mutation have been previously described *^26^*. C57BL/6J mice were purchased from the Jackson Laboratories (Bar Harbor, ME). All animal experiments were approved by the Animal Care and Use Committees of Vanderbilt University and University of Virginia. MSCV-MLL-AF9-IRES-GFP vector was provided by Dr. Scott Armstrong (Memorial Sloan-Kettering Cancer Center, New York, NY) and MSCV-BCR/ABL- IRES-GFP vector was provided by Dr. Michael Engel (University of Utah, Salt Lake City, UT).

### Retrovirus production

Retroviral vectors were transfected into Phoenix ecotropic retroviral packaging cells using Lipofectamine 2000 according to the manufacturer’s instructions. Forty- eight hours after transfection, viral supernatants were collected, filtered, and stored at −80°C.

### Induction of myeloid leukemia in mice

The MSCV-MLL-AF9-IRES-GFP retrovirus was used to induce AML in mice. Bone marrow cells were flushed from the tibias and femurs of *Tgif1*^+/+^ or *Tgif1*^-/-^ mice and lineage marker-negative (Lin^-^) cells were enriched using the Mouse Lineage Cell Depletion kit (Miltenyi Biotec, San Diego, CA). Lin^-^ cells purified from *Tgif1*^+/+^ or *Tgif1*^-/-^ mice were then transduced using low-speed centrifugation, or spinoculation, with MSCV-MLL- AF9-IRES-GFP retrovirus in the presence of 8 μg/ml polybrene at 1350g × 45 min (32°C). Transduced *Tgif1*^+/+^ or *Tgif1*^-/-^ Lin^-^ cells were resuspended in phosphate-buffered saline (PBS) and 1 × 10^5^ cells were injected intravenously into sub-lethally irradiated (4.5 Gy) *Tgif1*^+/+^ or *Tgif1*^-/-^ recipient mice, respectively. Reconstitution of transduced cells in recipient mice was evaluated by monitoring GFP expression in flow cytometry (FACS) analysis at 1-2 week intervals following transplantation. Recipient mice were euthanized for analysis when they developed palpable splenomegaly or appeared ill. Bone marrow and spleen cells from leukemic mice were isolated and stored frozen for later use.

The MSCV-BCR/ABL-IRES-GFP retrovirus was used to induce CML in mice. Bone marrow cells were flushed from the tibias and femurs of *Tgif1*^+/+^, *Tgif1^+/-^*, or *Tgif1*^-/-^ mice and single-cell suspensions of Lin^-^ c-Kit^+^ cells were obtained using the Mouse Lineage Cell Depletion kit and CD117 microbeads (Miltenyi Biotec, San Diego, CA). Lin^-^ c-Kit^+^ cells were spinoculated with MSCV-BCR/ABL-IRES-GFP retrovirus in the presence of 8 μg/ml polybrene for 45 minutes at 1350g (32°C). Transduced cells were resuspended in PBS and 8 × 10^4^ cells were injected intravenously into lethally or sub-lethally irradiated C57BL/6J recipient mice. Reconstitution of transduced cells in recipient mice was evaluated by FACS analysis of GFP expression two weeks after transplantation. Mice were monitored for development of disease as described above, at which time they were euthanized.

### Quantification of leukemia-initiating cell frequency

Leukemia cells from spleens of three *Tgif1*^+/+^ or three *Tgif1*^-/-^ mice with AML induced with the MSCV-MLL-AF9-IRES-GFP retrovirus were pooled, and 1 × 10^6^, 1 × 10^5^, 1 × 10^4^, 1 × 10^3^, 100, and 50 cells were transplanted into sub-lethally irradiated recipients. Animals were euthanized when they became visibly ill and the development of leukemia was confirmed. LIC frequency was determined by limiting dilution analysis with the ELDA (for Extreme Limiting Dilution Analysis) software package *^58^*.

### Chemotherapy studies in AML mice

For treatment studies, 2 × 10^3^ splenic leukemia cells from *Tgif1*^+/+^ or *Tgif1*^-/-^ mice were transplanted into sub-lethally irradiated recipients by tail vein injection. Two weeks after transplant, mice were treated by intraperitoneal injection with 100 mg/kg cytarabine once per day for five days and 3 mg/kg doxorubicin for three days. Mice were monitored closely for disease development and GFP expression was evaluated weekly by FACS analysis. Animals were euthanized when they appeared ill and transfer of disease was verified.

### RNA sequencing (RNA-seq) analysis of AML cells

RNA was extracted from sorted populations of GFP-expressing leukemia cells isolated from the bone marrow or spleen of *Tgif1*^+/+^ or *Tgif1*^-/-^ mice with AML (3 replicates each). RNA-seq and bioinformatics analysis was performed in the Vanderbilt Technologies for Advanced Genomics (or VANTAGE) Shared Resource. Sequence data were annotated with the University of California, Santa Cruz Genome Browser (https://genome.ucsc.edu/), and the TopHat and Cufflinks software packages were used for transcript alignment and quantification of gene expression. Gene set enrichment analysis *^59^* was carried out using the default weighted enrichment statistic to test whether differentially expressed genes (between *Tgif1*^+/+^ and *Tgif1*^-/-^ LSK cells) were randomly distributed or enriched at the top or bottom of the gene list. A false discovery rate of *≤* 0.25 was considered significant. Ingenuity Pathway Analysis (http://www.ingenuity.com/) was performed to group functionally related genes and assign them to biological pathways.

### RNA preparation and cDNA microarray analysis

Leukemic blasts from heparinized bone marrow or peripheral blood were enriched by Ficoll-Hypaque gradient centifugation, depleted of T lymphocytes and monocytes using magnetic beads (Dynal Biotech, Brown Deer, WI) conjugated with monoclonal antibodies to CD2 and CD14, respectively, and immediately cryopreserved. In the case of myelomonocytic (FAB M4) or monocytic (M5) leukemias, only the CD2 antibody was used. All cell populations contained at least 95% blasts and had an initial viability of ≥90%. Total cellular RNA was prepared from thawed cells on silica-gel membranes with on-column DNAse treatment (RNeasy Total RNA Isolation Kit, Qiagen, Valencia, CA). RNA integrity was assessed with an Agilent Bioanalyzer (Agilent Technologies, Palo Alto, CA), and samples showing evidence of degradation were discarded. Fluorescent labeling of probes was carried out in the Microarray Shared Resource of the Vanderbilt-Ingram Cancer Center, with incorporation of amino-allyl dUTP during reverse transcription and reaction of the amine-modified cDNA with monofunctional forms of Cy3 and Cy5 dyes. Dye-labeled cDNAs were then hybridized to glass slides that had been robotically spotted with an 11,000-element human cDNA library, and fluorescence intensity was measured with an AXON or GSI Lumonics dual laser scanner. Leukemic cell cDNAs were competitively hybridized with cDNA generated from a commercial reference RNA (Universal Human Reference RNA, Stratagene, La Jolla, CA) derived from a mixture of ten human cell lines. The ratio of transcript abundance in experimental samples to the RNA reference was first transformed by log_2_. Normalization of hybridization signals was then carried out with GeneTraffic software (Iobion, La Jolla, CA) using the following parameters: signal to background intensity <1.3, Cy3 intensity <150, and Cy5 intensity <200.

## Data Availability

Data included in the manuscript

## Acknowledgements

This work was supported by National Institutes of Health grants K08 HL089903 (to R.H.) and P30 CA068485 (to Vanderbilt-Ingram Cancer Center) and American Cancer Society Scholar Award RSG LIB-118462 (to R.H). We thank Drs. Ruud Delwel, Sanne Lugthart, and Bob Löwenberg for making available and analyzing microarray data from the Erasmus University study, Scott Boswell and Scott Armstrong for provision of materials, and Craig Jordan and Scott Hiebert for helpful discussions.

## Author Contributions

L.Y., J.A.M.-P., R.H. and S.J.B. conceived the studies. V.D.K., J.P.G., U.P.D. and D.W. provided study materials or patients. L.Y., J.A.M.-P., R.H., and D.M., collected and/or assembled data. L.Y., R.H., Y.S., J.P.G., U.P.D., M.J.K., D.W. and S.J.B. contributed to data analysis and interpretation. L.Y., J.A.M.-P., R.H. and S.J.B. drafted the manuscript. All authors gave final approval of the manuscript.

---

**Supplemental Table S1.** Patient characteristics.

| Characteristic | Number | Percentage |
| --- | --- | --- |
| Total number | 60 |  |
| Median age | 48 (range 3-79) |  |
| Age 3-17 | 6 | 10% |
| Age 18-45 | 21 | 35% |
| Age 46-60 | 19 | 32% |
| Age 60-79 | 14 | 23% |
| Males | 28 | 47% |
| Females | 32 | 53% |
| Newly diagnosed | 53 | 88% |
| Relapsed | 7 | 12% |
| Prior MDS | 21 | 35% |
| Cytogenetics |  |  |
| Good prognosis | 6 | 10% |
| Intermediate prognosis | 27 | 45% |
| Poor prognosis | 22 | 37% |
| Induction therapy | 49/53 newly diagnosed | 92% |
| Salvage therapy | 3/7 relapsed | 43% |
| Achieved CR | 35 | 58% |
| No response or PR | 17 | 28% |
| Remission not evaluated | 8 | 13% |
| SCT in CR1 or CR2 | 20 | 33% |
| Patients surviving | 17 | 25% |

**Supplemental Figure S1.**
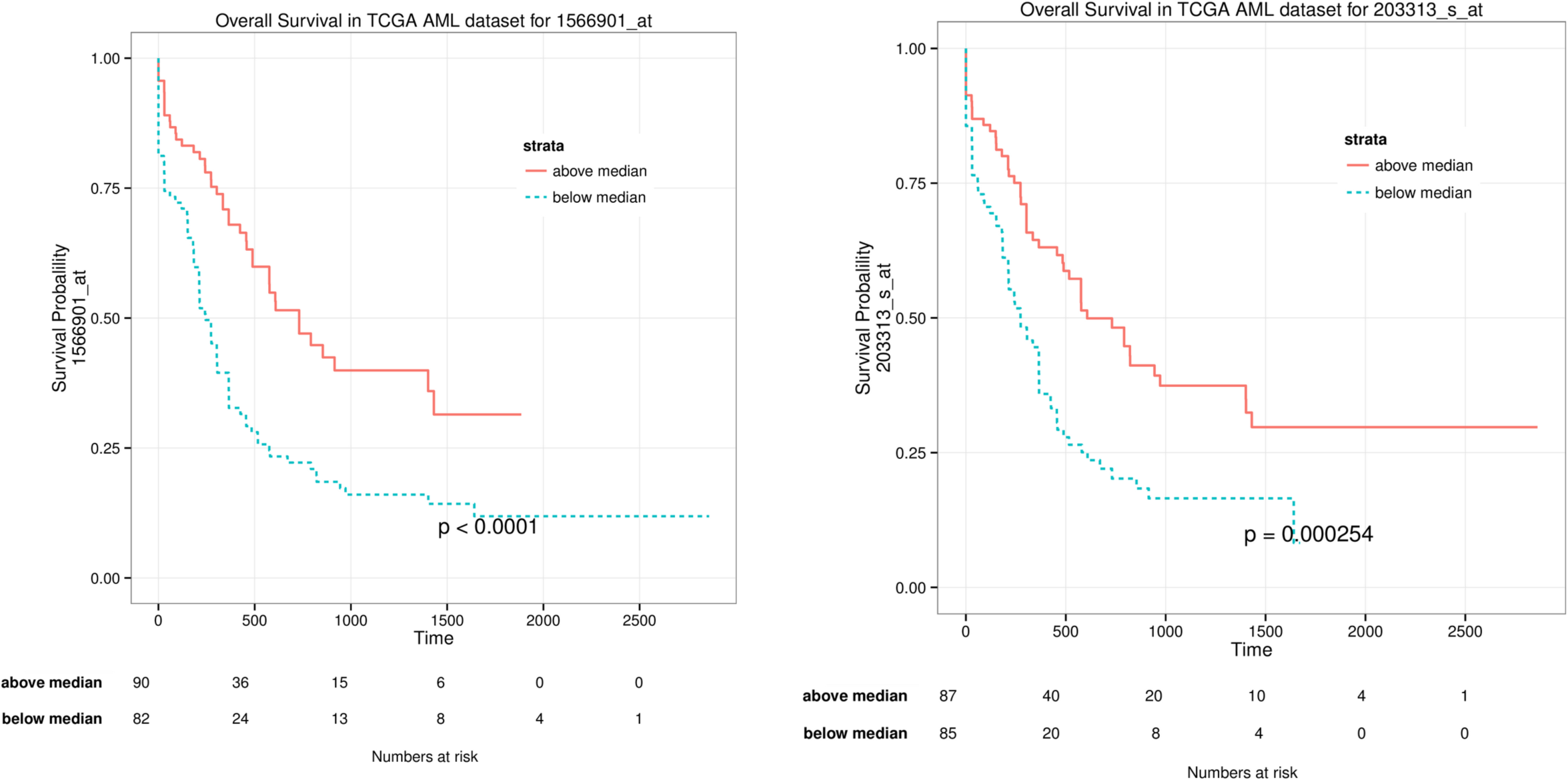
Kaplan-Meier analysis of OS as function of *TGIF1* expression: TCGA AML dataset. Kaplan-Meier analysis of OS for adult patients with AML from TCGA dataset, with *TGIF1* expression above the median (red continuous lines) or below the median (blue dotted lines) plotted for probe set 203313_s_at (*left*) and 1566901_at (*right*).

**Supplemental Table S2.** TGIF1 sequence variants in myeloblasts.

| Polymorphism | TGIF region | Previously reported | Number |
| --- | --- | --- | --- |
| -33C>A | 5' untranslated | Yes | 9/49 |
| -9G>A | 5' untranslated | No | 1/49 |
| 127C>G (P43A) | Coding | No | 1/49 |
| 420A>G (P140P) | Smad2-interacting | Yes | 7/49 |
| 487C>T (P163S) | Smad2-interacting | Yes | 7/49 |
| 488C>T (P163L) | Smad2-interacting | Yes | 7/49 |
| 713C>A (P238H) | Phosphorylation site | No | 1/49 |
| 723G>A (P241P) | Phosphorylation site | No | 1/49 |
| 744T>A (S248R) | mSin3A-interacting | No | 1/49 |

**Supplemental Figure S2.**
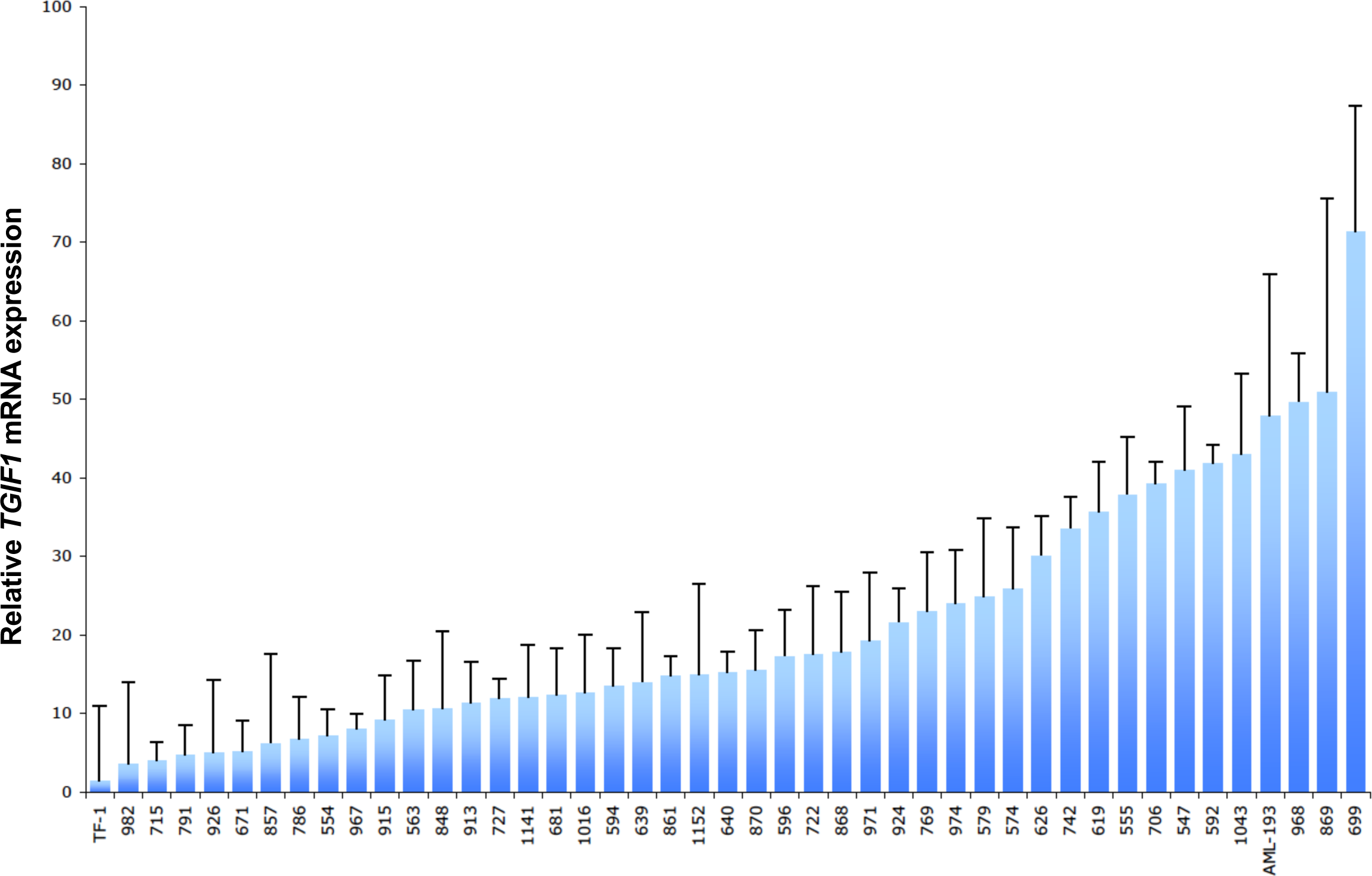
Quantitative RT-PCR analysis of *TGIF1* mRNA in leukemic blasts. Bar graph shows, in ascending order, mean *TGIF1* mRNA level ± SEM in myeloblasts from 43 patients with AML determined by real-time PCR analysis. *TGIF1* expression in human AML cell lines TF-1 and AML-193 are shown for comparison. The level of expression in the TF-1 cell line was assigned a value of 1 and all other measurements normalized to it.

**Supplemental Figure S3.**
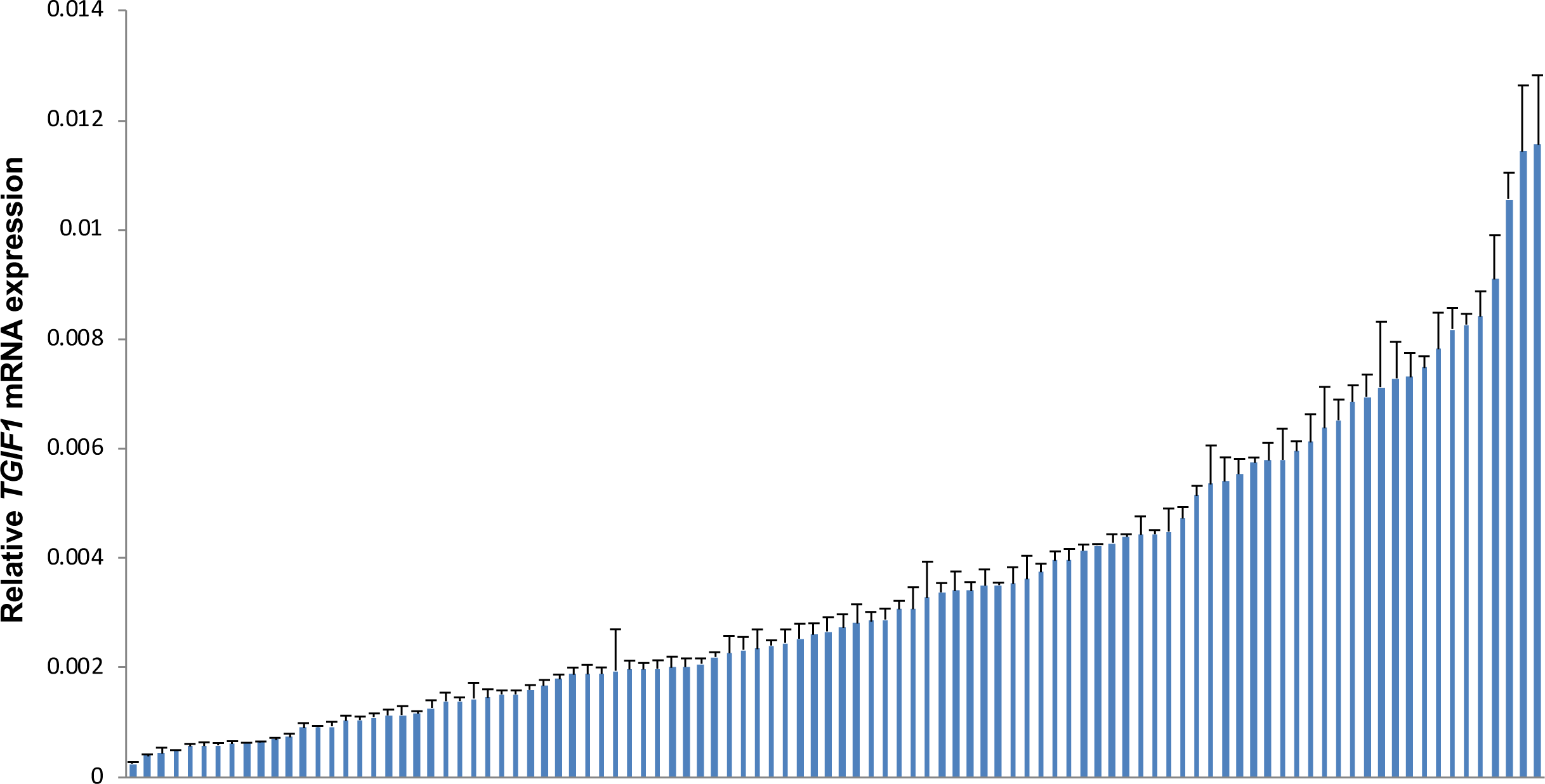
Quantitative RT-PCR analysis of *TGIF1* mRNA in immortalized lymphoblasts. Bar graph shows mean *TGIF1* mRNA level ± SEM in ascending order in immortalized lymphoblast lines derived from 100 normal individuals.

**Supplemental Figure S4.**
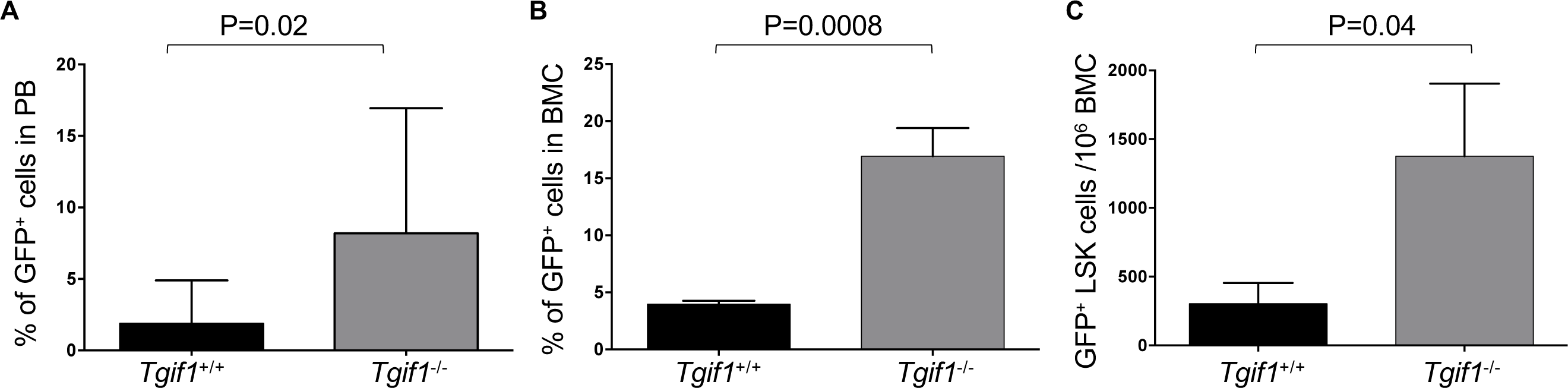
*Tgif1*^-/-^ mice with CML have more aggressive disease than in *Tgif1*^*+/+*^ mice. Enriched Lin^-^ c-Kit^+^ cells from *Tgif1*^+/+^ or *Tgif1*^-/-^ bone marrow cells were transduced with BCR-ABL-GFP retrovirus and transduced cells transplanted into lethally irradiated C57BL/6J recipients to induce CML. Six weeks post-transplant, flow cytometry was used to analyze **A**, percentage of GFP^+^ cells in PB; **B**, percentage of GFP^+^ cells in bone marrow; **C**, number of GFP^+^ LSK cells (or LSCs) in bone marrow.

**Supplemental Figure S5.**
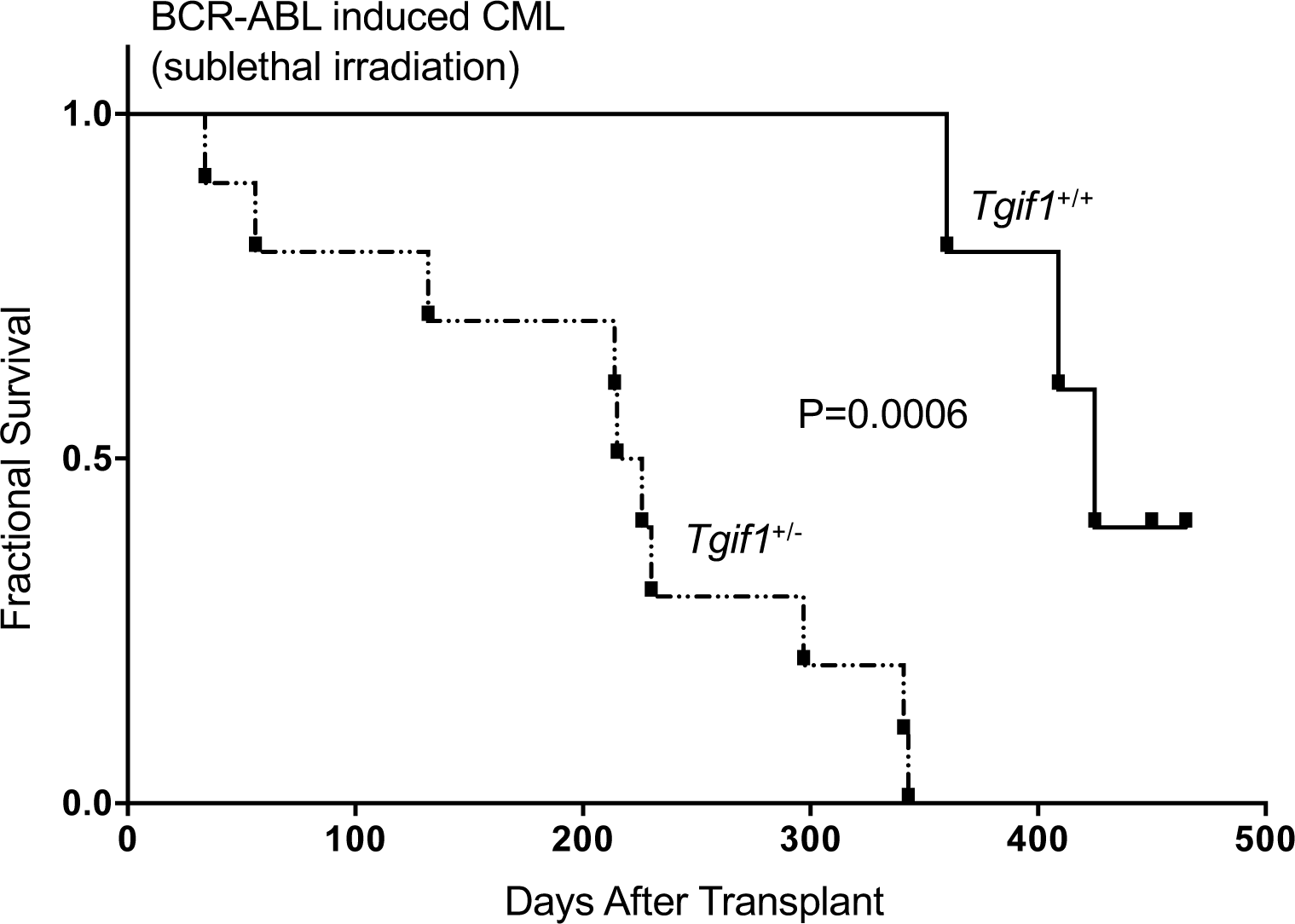
Kaplan-Meier analysis of survival in BCR-ABL-induced CML in mice as a function of *Tgif1* expression level. Enriched Lin^-^ c-Kit^+^ cells from *Tgif1*^+/+^ or *Tgif1^+/-^* bone marrow cells were transduced with BCR-ABL-GFP retrovirus and transduced cells transplanted into sub-lethally irradiated C57BL/6J recipients to induce CML.

**Supplemental Figure S6.**
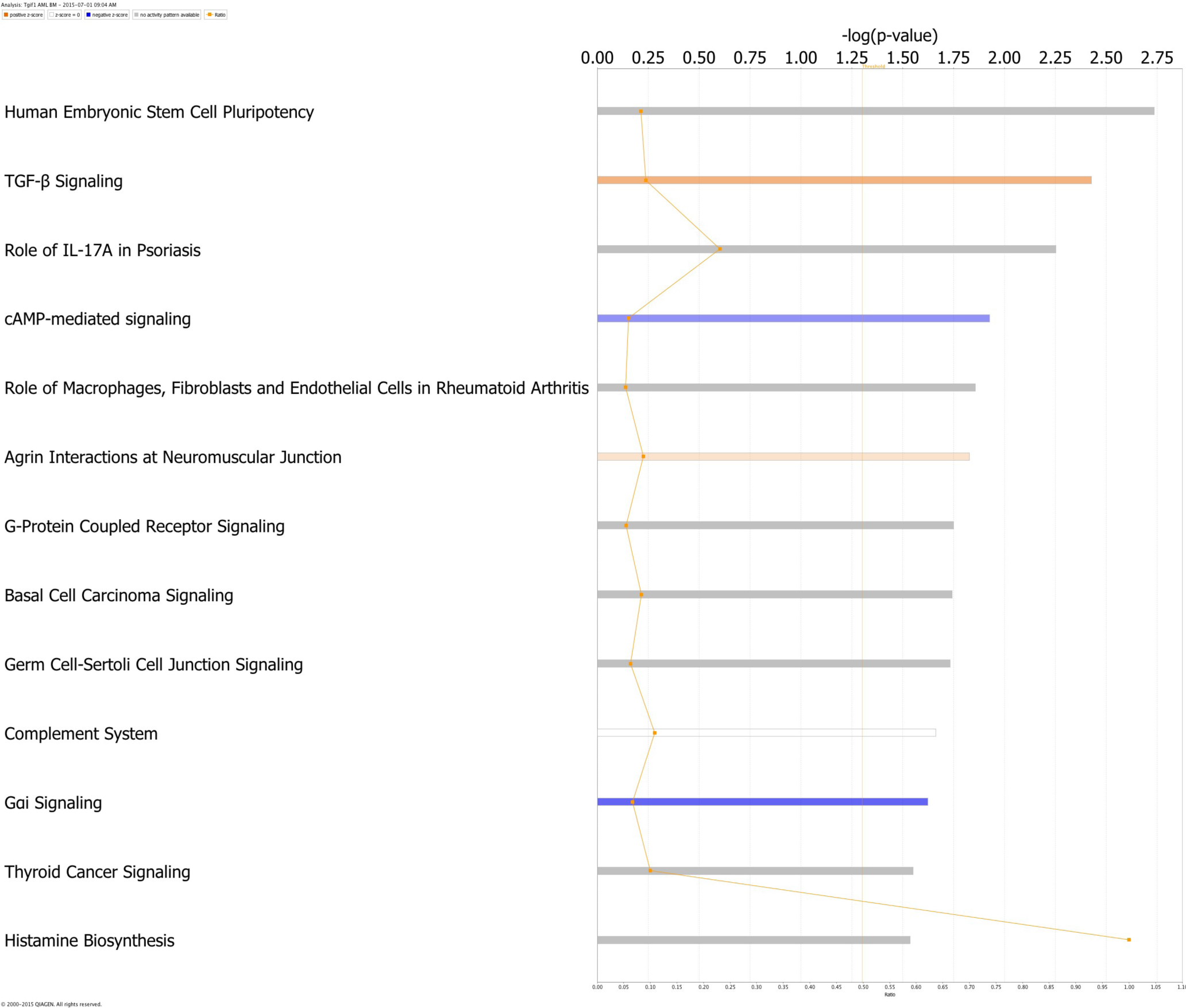
Canonical Pathway Analysis. Depicted are differentially utilized pathways in leukemia cells from *Tgif1*^-/-^ compared to *Tgif1*^+/+^ mice.

